# Exploring bed sensor sleep technology: insights from an interdisciplinary team in a geriatric assessment inpatient setting

**DOI:** 10.1101/2025.02.07.25321857

**Authors:** Yong Zhao, Cromwell G. Acosta, Yayan Ye, Karen Lok Yi Wong, Joanna Lawrence, Michelle Towell, Heather D’Oyley, Marion Mackay-Dunn, Bryan Chow, Lillian Hung

## Abstract

Sleep quality is a critical component of health and recovery for hospitalized older adults, yet current monitoring practices often lack the precision and detail required for effective intervention. This qualitative study aimed to evaluate the feasibility and acceptance of implementing the Sleepsense bed sensor for sleep monitoring in a geriatric inpatient hospital setting. This qualitative study involved interviews with 22 patients and focus groups with 33 interdisciplinary staff members. Data were analyzed using an interpretive description approach, guided by the technology acceptance model and unified theory of acceptance and use of technology, to explore the feasibility and acceptance of Sleepsense bed sensors in a geriatric inpatient setting. Key findings from thematic analysis emerged in three main themes representing the feasibility and acceptance of Sleepsense bed sensors among hospitalized older adults: user acceptance, integration with somnolog into clinical practice, and implementation barriers and practical challenges. Staff reported high acceptance of Sleepsense technology due to its nonintrusiveness and ability to reduce disruptive nighttime checks. However, challenges such as the need for consent, data interpretation and occasional inaccuracies were also identified. Integrating Sleepsense with existing care practices was recommended to enhance patient care while maintaining staff confidence. These findings underscore the potential of advanced sleep monitoring technologies in subacute care settings and highlight the importance of addressing implementation barriers for effective adoption.

**Author summary:** Sleep is crucial for hospitalized older adults, but traditional monitoring methods, such as nighttime checks, can be disruptive and may not always provide accurate assessments. We wanted to find out if the Sleepsense bed sensor would be acceptable to patients and staff in a hospital setting. We interviewed 22 hospitalized older adults and held group discussions with 33 staff members from various departments. Patients and staff valued the sensor’s ability to monitor sleep without disturbing rest, and staff recognized its potential to reduce nighttime checks. However, they emphasized that routine checks provide unique benefits, such as observing patient behavior and delivering personalized care, which the sensor alone cannot replace. We found that staff suggested integrating the sensor with existing practices to combine technological benefits with human oversight. They also identified challenges, including obtaining patient consent, interpreting sensor data, and managing occasional reading errors. We believe addressing these concerns through staff training and improving data reliability could support the sensor’s adoption in hospitals. By combining the strengths of technology with hands-on care, we see the Sleepsense bed sensor as a valuable tool to enhance sleep monitoring and improve care for older adults in hospital settings.

## Introduction

Sleep is a fundamental component of health, particularly for older adults, whose sleep patterns often become significantly disrupted due to advancing age, disease, interventions, or environmental disruptions (1,2). Furthermore, hospitalized older adults face additional challenges, such as noise, light exposure, pain, or frequent urges to micturate (3). These factors exacerbate sleep disturbances, hindering recovery and prolonging hospital stays. Insufficient sleep in this demographic increases the risk of chronic illnesses, such as heart and lung diseases, diabetes, and cognitive impairment (4). Additionally, poor sleep quality can weaken immune system efficiency, increasing the vulnerability of older adults to infections, delirium, uncontrolled pain, dehydration, and polypharmacy. These conditions contribute to prolonged institutionalization and functional decline (3,5). These disruptions underscore the importance of reliable sleep monitoring in hospital settings.

Sleep monitoring in older adults plays a vital role in detecting disorders such as sleep apnea, insomnia, and restless leg syndrome (6). These conditions are known to negatively affect physical health, mental functioning, and emotional stability, underscoring the importance of reliable assessment tools (7,8). Early identification of sleep disturbances allows targeted treatments, such as cognitive‒behavioral therapy for managing insomnia and the use of continuous positive airway pressure (CPAP) devices for sleep apnea, which can significantly enhance sleep quality and health outcomes (9). Moreover, consistent tracking of sleep patterns provides valuable data for evaluating the success of interventions and making necessary adjustments, thereby promoting sleep-facilitated recovery, enhancing physiological reserve, and perpetuating long-term sleep health with improved overall quality of life (QOL) as an outcome (10,11).

Innovations in artificial intelligence (AI) have introduced advanced solutions for addressing these challenges. The integration of AI with digital monitoring technology offers innovative tools for analyzing sleep characteristics, enabling clinicians to improve diagnosis and treatment (12,13). AI-powered sleep monitoring devices have the potential to support patient safety and quality of care with tailored treatment strategies to enhance individual health outcomes. Our study examined the Sleepsense bed sensor, a nonwearable AI-powered device, which provides real-time monitoring of sleep behaviors and vital signs, enabling healthcare providers to detect potential health issues early and tailor care strategies accordingly (14). This device uses passive sensing technology based on ballistocardiogram (BCG) signals detected through a force transducer, enabling noninvasive real-time sleep monitoring (14). Positioned under a bed’s wheel or leg, it captures cardiorespiratory and subtle body movements, providing unobtrusive data for healthcare applications (14).

The adoption of AI-powered technologies relies on optimal functionality and the willingness and preparedness of healthcare teams to integrate them into practice. Understanding healthcare professionals’ perspectives—including their concerns, perceived benefits, and practical challenges—is crucial to developing strategies that promote acceptance and effective use. This paper reports the experiences of patients and staff in implementing Sleepsense in a geriatric hospital unit. This study aims to examine the feasibility and acceptance of implementing the Sleepsense bed sensor for sleep monitoring in a geriatric care setting.

### Research question

What are the experiences of patients and interdisciplinary staff regarding using Sleepsense bed sensors for older adults in hospital units?

## Methods

### Design

Using the interpretive description approach (15), we conducted interviews involving 22 patients and focus groups involving 33 interdisciplinary staff members, including physicians, nurses, care aides, and other healthcare allies in a geriatric assessment inpatient unit. This qualitative exploratory design focuses on gathering practical insights from participants to inform tailored strategies for adoption. Interpretive descriptions are well suited for exploring the nuanced, complex experiences of staff in clinical contexts. This design allows us to yield practical, applied findings that can be used to refine the sensor design, improve training, and address specific concerns raised by staff, ensuring that the technology is more acceptable and feasible in real-world settings. The focus group method was chosen for staff participants because it allows for interactive discussions among participants, enabling the exploration of diverse viewpoints, experiences, and insights related to the research topic (16). We report the results following the consolidated criteria for reporting qualitative research (COREQ) (17) (see S1 Table for the COREQ checklist).

### Setting

This research was conducted at the Short-Term Assessment and Treatment (STAT) Centre at the University of British Columbia Hospital. The STAT Centre is a specialized hospital unit for older adults experiencing subacute health challenges that impair their ability to function independently. The unit provides comprehensive assessments, targeted treatments, and multidisciplinary care, addressing physical, cognitive, and psychological needs. Collaboration among healthcare professionals—including physicians, nurses, occupational, physiotherapistic and recreational therapists, and social workers—ensures holistic, individualized care for patients. The STAT Centre’s focus on innovation and patient-centered care made it an appropriate environment for evaluating the implementation of advanced technologies, such as the SleepSense bed sensor.

### Participants

The study employed convenience sampling to recruit patients and interdisciplinary staff from the geriatric unit, offering a practical and cost-effective approach to data collection. The sample size was adequate to address the research questions comprehensively. Recruitment efforts were facilitated through the placement of posters and flyers in staff conference rooms and on unit noticeboards. The eligibility criteria for staff participants included (1) employment on either a full-time or part-time basis, (2) direct involvement in patient care, and (3) willingness to participate in focus group discussions about the adoption of SleepSense bed sensor technology.

Patient participants were eligible if they (1) were admitted to the STAT Centre, (2) were capable of communicating and completing interviews, and (3) expressed a willingness to participate in the SleepSense bed sensor project. All participants completed the study, with no refusals or dropouts recorded.

### Sleepsense bed sensor placement and measurement

After consent was obtained from patients, the Sleepsense bed sensors were installed beneath one wheel of each patient’s bed, where they remained for a period of three months. The sensors collect data on sleep metrics, heart rate, and respiratory rate and generate alerts when necessary, which are remotely communicated through the viewer access platform accessible to all internet-connected workstations on wheels (WOW). The industry partner provided training to staff participants to familiarize them with the sensor’s functionality, including physical setup and removal, admitting and discharging patients from the assigned sensors, and, most importantly, data interpretation. This training enabled staff to incorporate sleep-related information into their clinical assessments, treatment plans, and overall care planning. After the three-month observation period, the Sleepsense bed sensors were removed from the wheels of the patients’ beds.

### Data collection

The conversational interviews for patients and focus groups for staff were led by the primary investigator, an Asian male nurse, C.A., who holds a master of science in nursing degree alongside his coinvestigators J.L. and M.T. The investigators conducted the in-person conversational interviews with patients at bedsides and focus groups with staff while participants were at the workplace. The interviews lasted approximately 10 minutes, and the focus groups lasted an average of 30 minutes, which were audio recorded and professionally transcribed verbatim. Field notes were also utilized during the group sessions to document nonverbal cues and other contextual factors that might have impacted the conversation. We used an interview guide with three questions:

1. How well do you think the Sleepsense bed sensor accurately reflects your sleep patterns and quality? (after showing Sleepsense data and explaining the sleep metrics to patients)
2. Did the Sleepsense bed sensor affect your sleep in any way?
3. Would you feel comfortable using the Sleepsense bed sensor again?

We used a focus group guide with four questions:

1. Is it feasible to use bed sensors to collect data on patients’ sleep patterns and quality?
2. How does Sleepsense data compare to the information you obtain from visual checks or somnologs?
3. What challenges have you encountered from adding Sleepsense to your workflow?
4. Do you think the data from the bed sensor are relevant or helpful in the overall planning of patient care?

### Analysis

Our iterative data analysis was informed by the Technology Acceptance Model (TAM) and the Unified Theory of Acceptance and Use of Technology (UTAUT), two widely recognized frameworks for evaluating user acceptance and adoption of technology (18,19). TAM emphasizes two core factors that influence technology acceptance: perceived usefulness and ease of use, whereas UTAUT expands on this by including four additional determinants: performance expectancy, effort expectancy, social influence, and facilitating conditions. Braun and Clarke’s six-step thematic analysis was applied to identify codes, themes, and insights regarding patients’ and interdisciplinary team members’ perspectives on adopting the Sleepsense bed sensor for patient sleep monitoring (20). The research team included nurses (L.H., C.A., and Y.Y.), J.L., a physiotherapist, M.T., an occupational therapist, a physician (Y.Z.), and a social worker (K.W.).

Y.Y. is an Asian female graduate with a degree in the Science of Nursing, K.W. is an Asian female PhD graduate student in social work, M.T. is an occupational therapist who also holds an MScOT degree, and Y.Z. is an Asian male graduate in Health Leadership and Policy with an M.D. degree. To ensure a thorough understanding of the data, all team members independently conducted multiple readings of the transcripts. After familiarizing themselves with the data, C.A. and Y.Z. performed the initial coding, focusing on elements relevant to the research questions. Data management, including transcripts, quotations, and codes, was facilitated via NVivo version 14.0. The initial codes were discussed and grouped into themes by C.A. and Y.Z. on the basis of observed similarities and frequencies. These themes were reviewed and refined collaboratively with all team members. Discrepancies or alternative interpretations were resolved through team discussions until a consensus was reached. Once the final set of themes and subthemes was established, C.A. and Y.Z. named and defined them to accurately reflect the data’s content, ensuring alignment with the research objectives. These themes were then interpreted in the context of the study’s aims. The initial draft was written collaboratively by C.A., Y.Y., K.W., and Y.Z., with all the authors contributing to the revisions and approving the final version. L.H. provided mentorship and guidance to trainee authors throughout the data analysis process.

### Ethical considerations

The study received ethical approval from the Behavioral Research Ethics Board at the University of British Columbia, Office of Research Ethics (H23-01577). All participants provided written informed consent after being fully briefed on the study’s purpose, details, potential benefits, associated risks, and their right to withdraw at any time. To safeguard confidentiality, pseudonyms were assigned to anonymize participant identities.

### Rigor

In this research, rigor was upheld through multiple strategies. Reflective conversations during weekly research meetings and the maintenance of reflexivity notes were key practices to ensure a thoughtful and consistent approach. Transferability was addressed by providing rich, detailed descriptions of the study context, participants, and process, enabling readers to assess the applicability of the findings to other settings. Investigator triangulation was applied by involving three diverse researchers in decision-making processes related to coding, analysis, and interpretation. Comprehensive documentation of the research process, including data collection and analysis, was meticulously maintained to track progress and establish an audit trail, promoting both accountability and transparency throughout the study. To enhance the trustworthiness of interpretations, direct quotations from interdisciplinary participants were included. These combined efforts demonstrate a robust and systematic approach to maintaining research rigor.

## Results

The study included 55 participants, comprising 22 patients and 33 staff members (see Table 1). Among the staff, nurses (45.4%) and care aides (27.3%) represented the largest groups, followed by physicians (21.2%)—mainly psychiatrists—a social worker and a total activity worker (TAW). The majority of staff were female (90.9%), with ages ranging broadly. The largest age group was 40–50 years (30.0%), followed by 30–40 years (27.3%), and both the 50– 60 and 60–70 age groups accounted for 18.2% each. Most staff identified as Asian (72.7%), and their work experience varied, with the largest proportion having 20–29 years of experience (39.4%), followed by 5–9 years (24.2%) and 10–19 years (24.2%). This diverse professional and demographic background provided a rich perspective on the implementation of the Sleepsense bed sensor. Among the patients, the gender distribution was predominantly female (63.6%), with ages ranging from 66--91 years. The largest age group was 65–79 years (54.4%), followed by 80–89 years (40.9%). The majority of patients were White (72.7%).

**Table 1.**
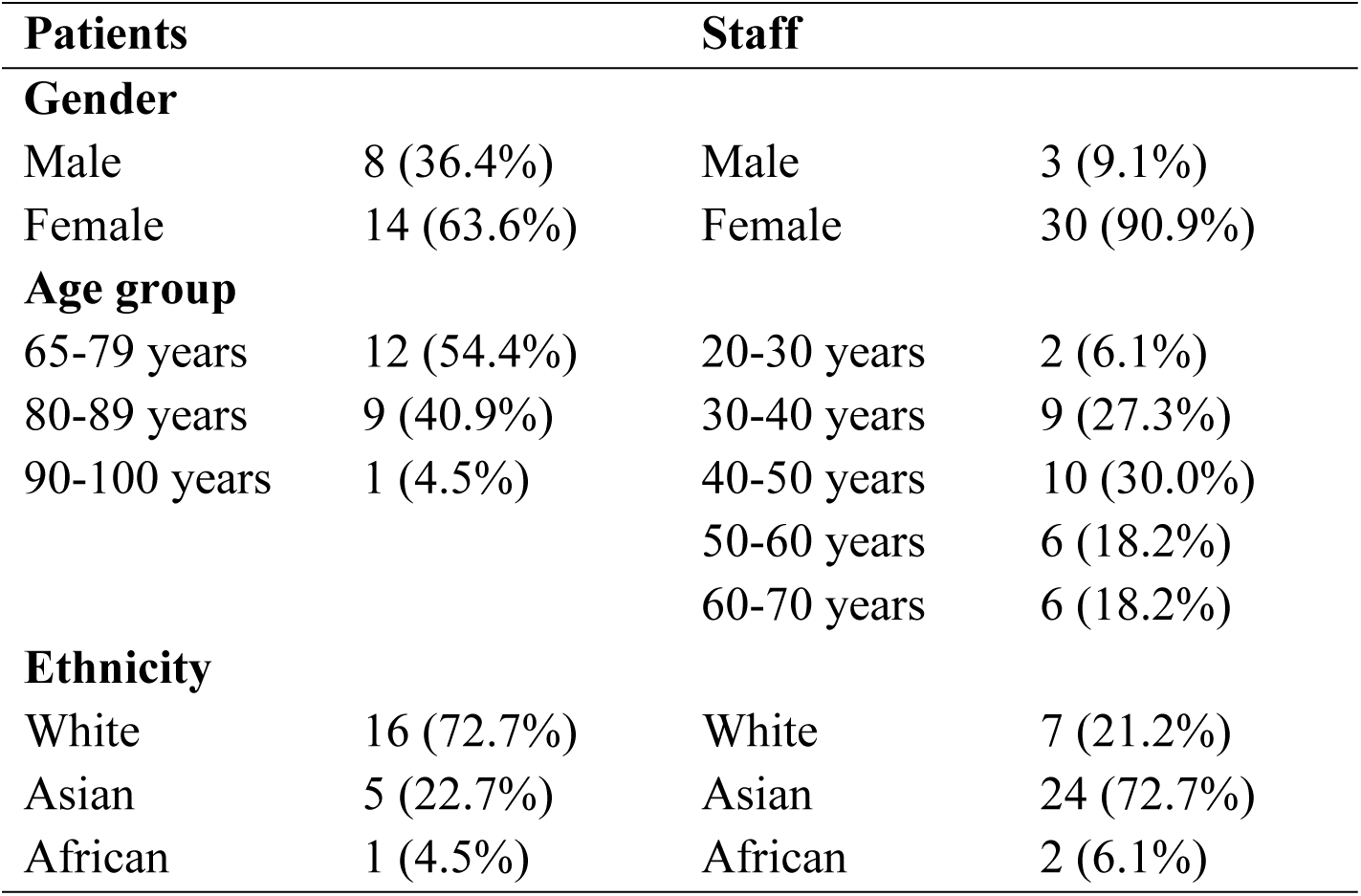
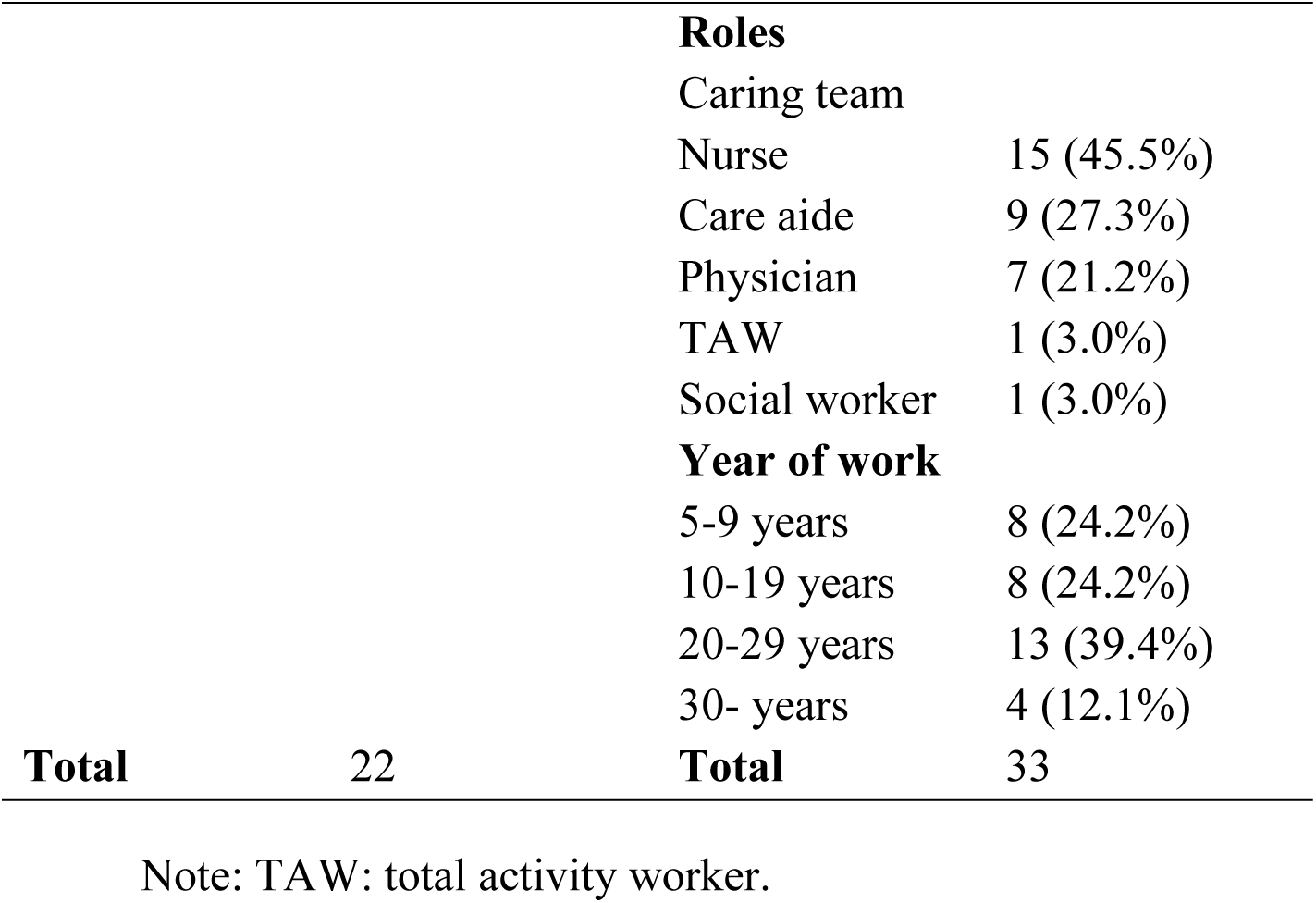
Demographic characteristics of the participants (n=55).

| Patients |  | Staff |  |
| --- | --- | --- | --- |
| **Gender** |  |  |  |
| Male | 8 (36.4%) | Male | 3 (9.1%) |
| Female | 14 (63.6%) | Female | 30 (90.9%) |
| **Age group** |  |  |  |
| 65-79 years | 12 (54.4%) | 20-30 years | 2 (6.1%) |
| 80-89 years | 9 (40.9%) | 30-40 years | 9 (27.3%) |
| 90-100 years | 1 (4.5%) | 40-50 years | 10 (30.0%) |
|  |  | 50-60 years | 6 (18.2%) |
|  |  | 60-70 years | 6 (18.2%) |
| **Ethnicity** |  |  |  |
| White | 16 (72.7%) | White | 7 (21.2%) |
| Asian | 5 (22.7%) | Asian | 24 (72.7%) |
| African | 1 (4.5%) | African | 2 (6.1%) |

|  |  | **Roles** |  |
| --- | --- | --- | --- |
|  |  | Caring team |  |
|  |  | Nurse | 15 (45.5%) |
|  |  | Care aide | 9 (27.3%) |
|  |  | Physician | 7 (21.2%) |
|  |  | TAW | 1 (3.0%) |
|  |  | Social worker | 1 (3.0%) |
|  |  | **Year of work** |  |
|  |  | 5-9 years | 8 (24.2%) |
|  |  | 10-19 years | 8 (24.2%) |
|  |  | 20-29 years | 13 (39.4%) |
|  |  | 30- years | 4 (12.1%) |
| **Total** | 22 | **Total** | 33 |

Note: TAW: total activity worker.

All the participants reported a high level of acceptance of this new sleep monitoring strategy in their daily clinical practice. Acceptance was reflected in their positive attitudes toward the sensor’s user-friendliness and perceived benefits. Notably, they viewed Sleepsense as an enhancement rather than a replacement for existing methods such as the somnolog sleep assessment. This preference highlights the feasibility of integrating Sleepsense into current practices, particularly when addressing patient needs and clinical workflow efficiency. The participants also identified various barriers and provided suggestions for improvement. The research generated three key themes: user acceptance, integration with somnolog into clinical practice, and implementation barriers and practical challenges (see Table 2).

**Table 2.**
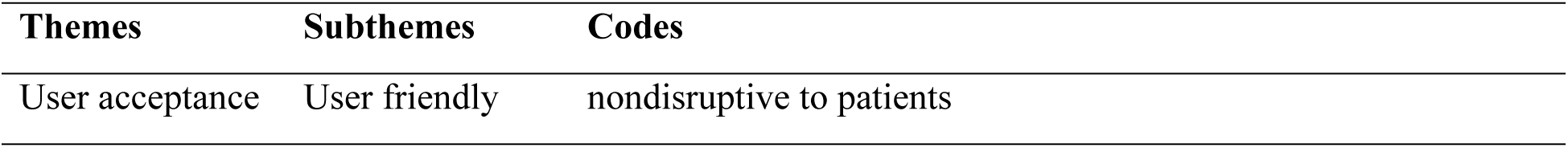
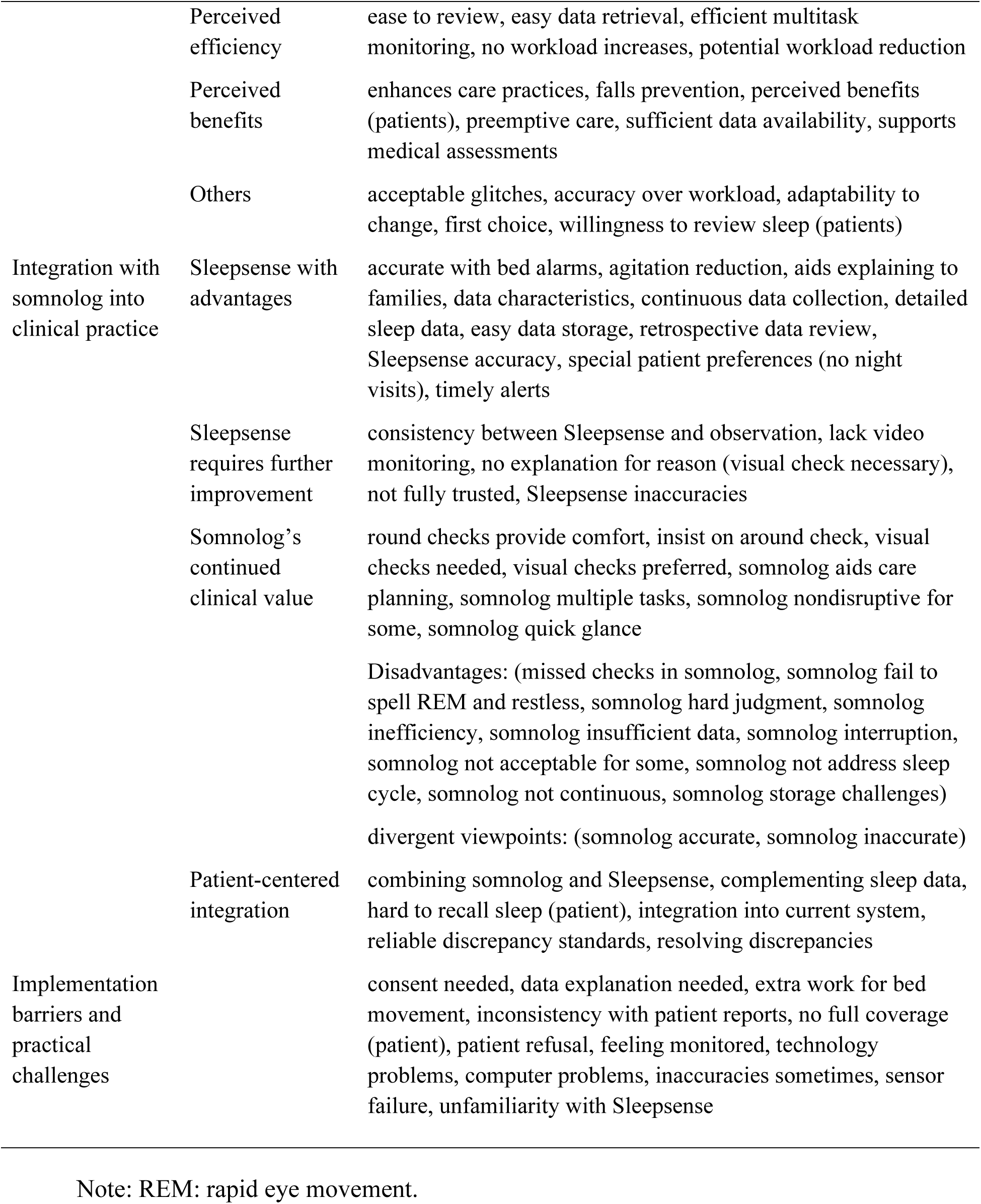
Codes and themes.

### Theme 1: User acceptance

#### Subtheme 1: User friendliness

All patient participants were provided with a review of their sleep data, which included results from the somnolog and bed sensor monitoring. All patients reported that the bed sensor did not cause any interruptions in their sleep. One patient, Issac, remarked, *“I didn’t even notice until you pointed it out! I was just thinking today—where did you put it?”* This reflects the sensor’s feasibility for long-term use without negatively impacting the patient’s sleep experience. Similarly, staff appreciated the sensor’s unobtrusiveness, making it a user-friendly addition to care routines.

#### Subtheme 2: Perceived efficiency

Staff highly praised the efficiency of the Sleepsense bed sensor, highlighting its ease of use for retrospective reviews, quick data retrieval, multi-patient monitoring, and its potential to reduce workload rather than add to it.

“I think the nurses will get used to it. At first, sure, they’ll probably do the sleep sensor check and then their regular routine rounds. But if it proves to be more accurate over time, they might start relying on it more and eventually cut down on the rounds.” - Gianna, nurse.

“It’s actually the quickest way to get information, especially if you’ve been away for a week. You come back, and you’re about to do your rounds, but you have no idea how your patient’s sleep pattern was last night. So, you just flip it open, check the data, and there you go!” - Harper, nurse.

“It allows you to monitor multiple people at the same time—like having 17 sets of eyes instead of just one. It makes things so much more efficient because you’re able to keep track of everyone at once without having to physically check on each patient constantly.” -Layla, nurse.

Such feedback underscores the acceptance of Sleepsense as a tool that supports clinical decision-making without increasing workload.

#### Subtheme 3: Perceived benefits

Both patients and frontline staff found the Sleepsense bed sensor to be beneficial, with staff highlighting its ability to provide detailed data, support care planning, and help prevent potential falls. These perceived benefits align closely with acceptance, as they reinforce the sensor’s value in promoting patient safety and well-being.

Layla, a nurse, shared, “You have to take some time to look at the monitor, but if it triggers when someone is up, you can step in before a fall happens. With regular alarms, they don’t go off until the person is already off the bed, but I think Sleepsense picks it up way quicker.”

Aurora, another nurse, noted, “The bed sensor lets us see the patient live—like we can tell what they’re doing, where they are, or if they’re sleeping, all without actually being there.

That doesn’t mean we’d stop physically checking on them every 30 minutes, but it’s a huge help for those times when we don’t want to disturb them or risk escalating their behaviors.”

Elizabeth, a TAW, explained, “You know, if they’ve had bad sleep or not, it can really affect their mood. Like, if I notice they didn’t sleep well, I might change my plans for them— maybe skip an activity that day or adjust it for another time. So yeah, it really helps.”

In addition to the three main subthemes, other factors also highlight the acceptance of Sleepsense. Patients expressed interest in reviewing their sleep data and often asked for more details about their sleep patterns. Most staff members showed trust in the Sleepsense data and preferred referencing it when conflicts arose between somnolog records and patients’ self-reports. They also demonstrated readiness for this practice change, embracing the challenges and occasional glitches brought by this innovative technology. Moreover, the staff expressed a strong commitment to geriatric care. For example, Lily, a nurse, remarked, *“It’s not so much about how much workload it adds, you know? The real question is how accurate the data is—because that’s what helps us provide better medical interventions for the patient, right? Even if the workload feels heavy, you’d still think it’s worth it if it gives the doctor reliable data to prescribe the right medications.”*

“The sensors act as a preventative measure to reduce incidents. They alert you to specific activities, allowing you to attend to individuals right away. This way, you can assess whether what’s happening poses a risk, like a potential fall, and take action to prevent it before it occurs.” - Emma, Nurse

### Theme 2: Integration with somnolog into clinical practices

The participants expressed their acceptance of Sleepsense technology while acknowledging that the current sleep recording method, somnolog, still has value and merits for continued use in clinical practice. This highlights the need for patient-centered strategies to integrate Sleepsense with existing sleep monitoring routines. Such integration is expected to increase the accuracy of overall care and medical planning without increasing staff workload. To address this further, we expand this theme into four subthemes, as outlined below.

#### Subtheme 1: Sleepsense with advantages

Most staff emphasized the advantages of the Sleepsense bed sensor, particularly its detailed data quality, compared with the current somnolog methods. Key benefits included improved data accuracy, continuous data collection, detailed insights into sleep patterns, convenient data storage, and the ability to conduct retrospective reviews.

Nora, a nurse, explained, “Sleepsense does a much better job at capturing each awakening. With somnolog, it only records every 30 minutes, so there could be awakenings happening in between that we’d completely miss. I feel like that’s the biggest difference.”

Camila, another nurse, added, “It can actually detect if you’re in light sleep or already in REM, you know? Yeah, it’s definitely more detailed.”

In addition to its data-related benefits, staff noted how the Sleepsense bed sensor enhanced their daily care and treatment routines. It supports specific patient needs, such as avoiding night visits, reducing agitation, improving communication with families, and providing timely alarms that facilitate more efficient care. Ava, a care aide, shared her perspective:

“With Sleepsense, nurses can reduce the need for frequent check-ins every half hour. It gives us peace of mind to keep the door closed, knowing we can monitor the patient’s breathing and ensure they’re okay without causing any distress. This way, we avoid the risk of opening the door and potentially agitating them to the point of needing security.”

#### Subtheme 2: Sleepsense requires further improvement

Staff acknowledged that while Sleepsense has many advantages, there is still room for improvement in its performance. For example, two recording errors occurred during the three-month study, and the system was unable to identify the reasons for awakenings, which has been regarded as the most necessary information they wanted. Staff also expressed a desire to incorporate video monitoring alongside the diagram data currently provided for a more comprehensive understanding of patient activity.

Both staff and patients mentioned that they have not yet fully trust Sleepsense yet. One female patient, Grace, shared, “It lets you know when you get up or lie down, and even if someone else—like a guest—sits on your bed during the day. Then, it’s like, ‘Oh, she had a three-hour nap today.’”

“This is more of a tool to help, not something that can replace nurses’ monitoring. I mean, unless you had a video setup where you could actually see the patient tossing and turning, like with a baby monitor.” - Isabella, nurse, PCC (patient care coordinator)

#### Subtheme 3: Somnolog’s continued clinical value

Compared with the Sleepsense bed sensor, the participants noted several disadvantages of the somnolog method, such as challenges with the implementation process (e.g., potential sleep interruptions—although not in all cases, inefficiency, and the lack of continuous monitoring).

Other issues included limited storage space and the system’s failure to address sleep patterns accurately or distinguish between REM and restlessness. Despite these drawbacks, participants acknowledged the value that somnologs bring to their practice. The key benefits included the comfort provided by routine round checks, the ability to quickly glance at data, multitasking capabilities, and its contribution to care plan adjustments.

Hazel, a care aide, shared, “I still want to finish my shift with my rounds, you know? Just to feel comfortable that our patients are still sleeping, okay, and that they’re in bed and doing fine.”

Amelia, a psychiatrist, added: “What I like about somnolog is the extra details, like ‘up and restless,’ ‘up and incontinent,’ or ‘up and calm,’ which we don’t truly get with Sleepsense. What I’d be worried about is if we move to a system where there aren’t any visuals or checks to see what’s happening with people overnight.”

Ellie, a nurse, mentioned, “I truly like the overall view that somnolog gives us—it’s handy for rounds. Instead of having to go through each patient and scroll, I definitely prefer that quick glance.”

#### Subtheme 4: Patient-centered integration

When discrepancies arose between patients’ self-reports, staff inspection, and Sleepsense bed sensor monitoring, participants (patients and staff) generally chose to trust the objective monitoring data from the sensor. Many patients admitted that they could not accurately recall or report their sleep from the previous night. However, discrepancies still exist between other patients’ reports and staff observations. For example, Nurse Isabella shared, *“If the patient’s a pill seeker, they’ll always be like, ‘I’m not sleeping, I’m not sleeping,’ and then the doctor just keeps ordering sleep meds. But if the sensor is showing, ‘Actually, you are sleeping,’ it changes everything.”* Similarly, care aide Scarlett noted a case where her initial assumption was incorrect: *“The patient told me, ‘I couldn’t sleep last night,’ but when I checked on her, she was in bed the whole time with her eyes closed, so I thought she had been sleeping all night. But then I looked at the graph, and it showed she was moving—just little movements, you know? Clearly, she wasn’t actually sleeping.”* These examples illustrate why most participants favored the objective data provided by Sleepsense, trusting its ability to capture insights beyond human observation or patient self-reports.

Patients also expressed great interest in their Sleepsense data, often asking to review their sleep reports and even requesting to see previous records in a sequence. This can strengthen engagement and trust in the implementation process. However, frontline staff emphasized that Sleepsense should complement, not replace, existing care routines. As Nurse Nova explained, *“I’d like a combination of all the options. It’s great to have the sensor for a detailed report on their sleep quality, like whether they’re getting deep sleep. But I also want to check in visually to make sure they’re breathing and doing okay. Plus, having their self-reports adds another layer, giving us a better overall picture of their health. So, a mix of all these would be ideal!”*

The staff appreciated Sleepsense’s continuous data collection and detailed metrics but valued somnolog’s quick-glance functionality and its role in care planning. This suggests that acceptance hinges on Sleepsense complementing, rather than disrupting, established practices. Additionally, some staff highlighted areas requiring improvement. These insights indicate that while Sleepsense is feasible for integration, optimizing its features is essential to fully meet clinical needs.

### Theme 3: Implementation barriers and practical challenges

The most frequently mentioned barrier was the issue of obtaining consent, which was reported mainly by physicians. Some patients were suspicious and refused to participate, whereas others, due to cognitive impairments, were unable to provide consent themselves. In these cases, their families often struggled to understand sleep monitoring technology and needed further explanation. One patient shared, “*I had the feeling that I was being watched and controlled*,” which might hinder acceptance.

Mila, a social worker, explained, “We have to make sure we get consent first, you know? We have to explain to them, ‘Oh, we’re using something that will monitor you while you’re asleep.’ And that might be one of the reasons why some patients choose not to participate.”

Lucas, a physician, added, “Sometimes patients disagree with what we observe on the sleep log, and it is hard to argue with them. My biggest concern would be individuals who are more suspicious or paranoid—one person, for example, didn’t really want to be monitored. But aside from that, it seems to be well tolerated.”

Staff also noted that during the initial stage of implementation, more work was required with the bed sensors. Since not all patients had Sleepsense monitoring, they had to place and remove the sensors for each bed, which added extra tasks. Nurse Isabella mentioned, “*One barrier is doing bed moves. If all the beds had sensors, we wouldn’t have problems—just transfer from one bed to another and clean. However, since not all of them have sensors…*”

In summary, both patients and staff accepted the Sleepsense bed sensor, with patients reporting that it did not disrupt their sleep. The staff praised the sensor’s efficiency, highlighting its ability to streamline data retrieval, monitor multiple patients at once, and support timely interventions to prevent falls. While patients expressed interest in reviewing their sleep data, staff emphasized that Sleepsense should complement, not replace, traditional care routines.

However, challenges, such as obtaining consent, sensor placement during bed moves, and occasional technological glitches, were noted. Despite these barriers, staff recognized the sensor’s value in enhancing care and preventing incidents, with the potential for future improvements in data accuracy and integration with existing systems.

## Discussion

This study shows good acceptance and feasibility of this innovative technology in a subacute hospital setting. The participants consisted of 55 individuals with 33 staff members in diverse roles and a significant representation of nurses, physicians, and care aides. The staff’s various professional experiences offered a robust foundation for evaluating the sensor’s acceptance and feasibility. Patients are predominantly female, older adults aged 65--89 years, and largely identified as White. This demographic composition emphasizes the relevance of the findings to populations commonly found in subacute care settings within the city where we conducted the study.

The findings highlighted strong acceptance of the Sleepsense bed sensor among staff and patients, who emphasized its efficiency, nonintrusiveness, and perceived benefits in enhancing care practices. Staff particularly valued its ability to provide detailed, continuous data, which facilitated timely interventions and reduced the need for disruptive physical checks. These benefits align with established strategies for improving sleep in older adults in acute care settings, such as noise reduction, minimizing nighttime interruptions, and assessing underlying sleep disorders (21). Together, these findings offer a robust foundation for integrating Sleepsense into clinical care.

A scoping review revealed that the sensor setup, whether unobtrusive or not, plays a role in determining acceptability as an implementation outcome (22). One feasibility study reported no complaints from participants regarding the use of an unobtrusive, contactless, portable bed sensor in a clinical rehabilitation stroke ward (23). This sensor, placed beneath the mattress, was designed to detect pressure. Another study conducted in home settings reported the feasibility and acceptance of a similar pressure-detecting bed sensor placed under the mattress, with no intentional dropouts from participants (24). However, some participants in that study mentioned behavioral changes, such as “intentionally decreasing trips to the bathroom,” owing to the constant awareness of being monitored. This sentiment was echoed by a few participants in our study, although most reported being largely unaware of the sensor’s presence. Furthermore, the discomfort caused by the sensor under the mattress led to sensor pad removal in Lach’s study. In contrast, this issue was not reported in our study, which may be attributed to the placement of the sensor under the bed leg rather than beneath the mattress. This positioning may have mitigated physical discomfort and contributed to sustained usage and acceptance within our research context.

However, barriers to implementation were also identified, including occasional inaccuracies, concerns about data interpretation, and the need for complementary video monitoring to contextualize recorded data. Our pilot study noted additional staff-perceived challenges, such as technology-related issues, the learning curve, privacy issues, and initial doubts about the reliability of Sleepsense (14). Over time, staff built confidence in the technology and expressed readiness to adopt it despite minor glitches. These insights underscore the importance of training and ongoing support to ensure successful adoption and integration into clinical workflows.

The current sleep assessment routines using somnologs have certain limitations, as reported by frontline staff during preintervention focus groups (14). These included occasional misses of “got-up” events during the checking intervals and overreporting due to challenges in judgment. Discrepancies between somnolog and Sleepsense were noted in this study, particularly when staff marked “awake but in bed,” whereas Sleepsense recorded the patient as “sleeping.” Focus group discussions revealed that staff sometimes struggled to assess sleep status confidently when patients kept their eyes closed and remained still. Similarly, differences arose in identifying “sleeping but restless” versus actual awakenings, underlining the need for Sleepsense objective data to increase staff confidence in their judgments. In some cases, staff resorted to further verifying somnolog data by flashing lights or asking questions before bed sensor application, which could disrupt patients’ sleep.

Despite these issues, somnolog provided valuable, detailed information that Sleepsense did not capture, such as events such as “bathroom,” “commode,” “urinal,” “medication,” “awake but in bed,” “sleeping but restless,” and “awake and out of bed.” In contrast, Sleepsense primarily recorded “got up” events. These granular data are crucial for care and medical teams in understanding patients’ sleep behaviors. Feedback from focus groups indicated that while staff acknowledged Sleepsense’s advantages, including its sensitivity and efficiency, they preferred to retain somnolog for its established clinical value and the reassurance it provided through direct visual observations. Staff appreciated how somnolog’s detailed records complemented the data from Sleepsense, particularly when making care decisions or adjusting medications. Integrating both systems into current workflows would increase the feasibility of Sleepsense implementation, allowing for a more comprehensive, accurate, and balanced approach to patient care.

The participants advocated combining Sleepsense with existing tools, such as somnolog, to create a dual approach that balances subjective and objective data collection in a multifaceted manner, thereby facilitating a more personalized patient care approach. This strategy acknowledges the strengths and limitations of Sleepsense bed sensor clinical integration while maintaining patient-centered care, which is supported by the findings of Landry et al. (1). Their study recommended the inclusion of both subjective and objective measures (e.g., the Pittsburgh Sleep Quality Index, Consensus Sleep Diary, and Actigraphy) as best practices for examining sleep quality in older adults. Overall, the findings emphasize the need for iterative technological improvements and strategies to foster trust and adaptability among clinicians. These strategies should be tailored to clinicians’ specific needs, ensuring the effective and sustainable use of Sleepsense technology—not limited to monitoring but, most importantly, providing sleep data that can guide clinical decision-making in the care of hospitalized older adults in a timely manner.

### Strengths and limitations

The strength of this study lies in its diverse participant pool, which includes older adults in subacute care settings and an interdisciplinary team of various healthcare professionals. This inclusivity allowed for multiple perspectives and comprehensive assessments, offering a nuanced understanding of the innovation’s impact on patient care and clinical workflows. However, the study focused exclusively on older adults in subacute care settings, which limits the generalizability of its results to other populations or care environments. Expanding research to include diverse patient groups and settings will be necessary to validate and extend the applicability of these findings.

### Conclusions

This study shows that the Sleepsense bed sensor is an acceptable and feasible tool in hospital inpatient settings, enhancing patients’ sleep health and overall well-being while providing detailed information and confidence to staff workflows. The integration of Sleepsense with existing sleep assessment tools has demonstrated the potential to provide a more comprehensive understanding of patients’ sleep patterns, supporting informed clinical decision-making. Successful implementation requires addressing barriers and providing thorough training for frontline staff, emphasizing the importance of support and usability. Future research should focus on long-term effects, broader participation, and the development of interdisciplinary strategies to support innovative practices that benefit both patients and healthcare providers.

## Ethical approval and consent to participate

The research was approved by the Behavioral Research Ethics Board at the University of British Columbia, Office of Research Ethics (H23-01577), on 8 September 2023. All participants provided written informed consent after being fully briefed on the study’s purpose, details, potential benefits, associated risks, and their right to withdraw at any time. To safeguard confidentiality, pseudonyms were assigned in the manuscript to anonymize participant identities.

## Consent for publication

Not applicable.

## Availability of data and materials

All the data generated or analyzed during this study are included in this published article and its supplementary information files.

## Authors’ contributions

L.H.: Conceptualization, Formal analysis, Writing—Review and editing, supervision, funding acquisition. Y.Z.: Formal analysis, data curation, writing—Original draft preparation. C.A.: Methodology, data curation, writing—review and editing, project administration, funding acquisition. Y.Y.: Formal analysis, data curation, writing—Original draft preparation. K.W.: Formal analysis, writing—Original draft preparation. J.L.: Writing—Review and editing. M.T.: Writing—Review and editing. H.D.: Writing—Review and editing. M.M.: Writing—Review and editing. B.C.: Writing—Review and editing. All the authors read and approved the final manuscript.

## Acknowledgments

Not applicable.

## Supporting information

S1 Table. Consolidated criteria for reporting qualitative studies (COREQ): 32-item checklist

